# Impact of the COVID-19 pandemic on the provision of routine childhood immunizations in Ontario, Canada

**DOI:** 10.1101/2021.05.11.21257048

**Authors:** Pierre-Philippe Piché-Renaud, Catherine Ji, Daniel S. Farrar, Jeremy N. Friedman, Michelle Science, Ian Kitai, Sharon Burey, Mark Feldman, Shaun K. Morris

## Abstract

**Background:** The COVID-19 pandemic has a worldwide impact on all health services, including childhood immunizations. In Canada, there is limited data to quantify and characterize this issue.

**Methods:** We conducted a descriptive, cross-sectional study by distributing online surveys to physicians across Ontario. The survey included three sections: provider characteristics, impact of COVID-19 on professional practice, and impact of COVID-19 on routine childhood immunization services. Multivariable logistic regression identified factors associated with modification of immunization services.

**Results:** A total of 475 respondents answered the survey from May 27^th^ to July 3^rd^ 2020, including 189 family physicians and 286 pediatricians. The median proportion of in-person visits reported by physicians before the pandemic was 99% and dropped to 18% during the first wave of the pandemic in Ontario. In total, 175 (44.6%) of the 392 respondents who usually provide vaccination to children acknowledged a negative impact caused by the pandemic on their immunization services, ranging from temporary closure of their practice (n=18; 4.6%) to postponement of vaccines in certain age groups (n=103; 26.3%). Pediatricians were more likely to experience a negative impact on their immunization services compared to family physicians (adjusted odds ratio [aOR]=2.64, 95% CI: 1.48-4.68), as well as early career physicians compared to their more senior colleagues (aOR=2.69, 95% CI: 1.30-5.56), whereas physicians from suburban settings were less impacted than physicians from urban settings (aOR=0.62, 95% CI: 0.39-0.99). The most frequently identified barriers to immunizations during the pandemic were parental concerns around COVID-19 (n=305; 77.8%), lack of personal protective equipment (PPE; n=123; 31.3%) and healthcare workers’ concerns of contracting COVID-19 (n=105; 26.8%).

**Conclusions:** COVID-19 has caused substantial modifications to pediatric immunization services across Ontario. Strategies to mitigate barriers to immunizations during the pandemic need to be implemented in order to avoid immunity gaps that could lead to an increase in vaccine preventable diseases.

**HIGHLIGHTS:**

- We have conducted a descriptive, cross-sectional study by distributing online surveys to pediatricians and family physicians across Ontario to assess the impact of the COVID-19 pandemic on their immunization practices.
- The COVID-19 pandemic has caused a substantial decrease in in-person visits and a related disruption to routine childhood immunization services during the first wave of the pandemic.
- The main barriers to immunizations during the pandemic included parents’ and healthcare providers’ concerns of contracting COVID-19, and lack of appropriate personal protective equipment (PPE).
- Solutions to maintain childhood immunizations during the pandemic included assistance in providing PPE to clinical practices, dedicated centers for vaccination, and parental education.

## BACKGROUND

The coronavirus disease 2019 (COVID-19) pandemic is causing an unprecedented impact on the delivery of healthcare services at all levels, including on routine childhood immunizations.^1-3^ Studies from France and the Centers for Disease Control (CDC) in the United States (US) have revealed declines of up to 70% in routine childhood vaccine coverage since the beginning of the COVID-19 pandemic.^4-6^ Disruptions to immunizations secondary to outbreaks of infectious diseases other than COVID-19 have previously resulted in increased rates of vaccine preventable diseases (VPDs), highlighting the current risks to Canadian children and their communities.^7,8^ Some low- and middle-income countries have also reported reduced access to mass vaccination programs and an increase in cases of measles attributed to COVID-19.^9,10^ In response to the pandemic, various public health and governmental institutions in Canada have recommended that routine immunizations constitute an essential health service that should not be deferred.^11,12^

In Canada, about half of the hospitalized children in Canada were admitted for reasons other than COVID-19, which was incidentally detected.^13^ Aside from cases of multisystem inflammatory syndrome in children (MIS-C) and its temporal association with COVID-19, which remains poorly understood, children and young adults are generally mildly affected by the disease when compared to adults.^14-16^ The indirect impacts of the COVID-19 pandemic on access to medical services, including routine immunizations, represent an important burden on children’s health that may outweigh the burden of the infection itself.^15,17,18^

In Ontario, school-based immunization of adolescents with hepatitis B vaccine (HBV), human papillomavirus (HPV) vaccine and meningococcal quadrivalent vaccine is delivered by public health units, but all other routine infant and childhood vaccines are most often administered in primary care clinics by family physicians and pediatricians. Some of the publicly funded vaccines can also be administered by pharmacists, but only for individuals older than five years old.^19^ While there are anecdotal reports of caregivers not bringing their children to primary care providers for immunizations and of clinicians that have ceased to provide these services due to perceived risk of COVID-19, there are currently limited data to quantify the magnitude of this issue and identify contributing factors. Furthermore, whether the COVID-19 pandemic has impacted access to immunization differentially for specific groups of children is not known. The scarcity of data on vaccination practices in Canada during the COVID-19 pandemic precludes any specific evidence-based recommendations regarding best practices in safely maintaining routine childhood immunization.^11,12,20,21^ The purpose of this study is to rapidly generate data to better understand the impact of COVID-19 on physicians’ immunization practices in Ontario.

## METHODS

This is a descriptive cross-sectional study consisting of a self-administered online survey. The survey was distributed electronically to pediatricians and family physicians across the province of Ontario. Emails were initially sent to a listserv of 347 pediatricians from the Greater Toronto Area (GTA) on May 27, 2020. An email was then sent on June 15, with reminders on June 22 and 29, to the Pediatricians Alliance of Ontario listserv, which comprises 1,313 pediatricians working in Ontario, including the 347 pediatricians from the GTA listserv.^22^ The survey was also distributed concomitantly on June 12 and 25 to family physicians using a listserv from the University of Toronto Department of Family and Community Medicine, which comprises 1,983 family physicians throughout the GTA and beyond. After having completed the survey, the respondents were given an individual link to another independent survey, where they could provide their name and email address to participate in an incentive draw for one of three $100 gift cards. The goal was to maximise participation and ensure adequate external validity and representation from the target population.^23^ Participants could decide not to opt in for the incentive draw if they wished to keep their responses completely anonymous. The collected emails and names were not linked with the participants’ responses to maintain confidentiality. Approval from The Hospital for Sick Children Research Ethics Board (REB) was obtained for this study (REB 1000070361).

### Survey Instrument

The survey instrument was developed with input from hospital and community-based physicians from general pediatrics, infectious diseases and family medicine. The survey incorporated previously published questionnaires on immunization practices of healthcare providers (HCP) and studies on HCP characteristics associated with up-to-date vaccination coverage of their patients.^24-27^ The survey was designed using Research Electronic Data Capture (REDCap) software (version 10.0.4; Vanderbilt University) and distributed by email as previously described.^28^ It was pilot tested with select pediatricians and family physicians from the target study population to ensure clarity of questions and instructions, ease of navigation and time for completion.^29^ The survey included three main sections: (1) HCP socio-demographic and practice characteristics, (2) impact of COVID-19 on clinical practice, and (3) impact of COVID-19 on immunization services.

Physician’s characteristics collected in the first section of the survey included physician’s specialty (family physician, general pediatrician or pediatric subspecialist), sex (male, female or prefer not to say), years in practice (<5 years, 5-20 years or >20 years) and country of medical training (Canada or outside Canada). Practice characteristics collected included setting (urban, suburban or rural), type of practice site (community solo practice, community group practice, family health team or hospital), practice forward sortation area (defined by the three first characters of the practice’s postal code), proportion of patients <19 years of age (for family physicians only), as well as availability of personal protective equipment (PPE) in physicians’ practices. These questions were selected based on previously published studies documenting HCP characteristics that may be associated with adequate on-time immunization coverage of their patients.^26,27^

The second section of the survey aimed to quantify the impact of the COVID-19 pandemic on physician’s clinical practice. Respondents were asked if the pandemic caused the physical closure of their practice, and if so, the specific reasons associated with closure. If their practice had remained open, respondents were asked to indicate the proportion of visits conducted in-person before the pandemic and during the pandemic.

The third section of the survey included questions aiming to assess the impact of COVID-19 on the provision of immunization services. Respondents were asked to select from a list of what they felt were significant barriers to their immunization services during the pandemic. The survey also included questions on respondents’ attitudes towards vaccines during the pandemic, such as if they felt it was preferable to postpone routine childhood immunizations to limit the spread of COVID-19 and risk for HCP. Answers for these questions were based on a five-point Likert scale (strongly agree, agree, neutral, disagree or strongly disagree). Participants were also asked if they provided vaccines to patients who were referred from other clinics, and how they managed their patients who have missed immunizations due to school closures. Lastly, the survey included two open-ended questions prompting respondents to describe any further barriers to the delivery of immunization services, as well as ideas and solutions on how routine childhood immunizations could be safely maintained during the COVID-19 pandemic. Physicians who reported not administering vaccines to children or teenagers in their regular practice, or family physicians who do not see children in their usual practice, were only asked the questions on attitudes regarding vaccines, as well as the open-ended question about potential solutions to maintain childhood immunization services. The detailed survey instrument can be found in the supplementary material (Appendix 1).

### Statistical Analysis

Survey responses were analysed using Microsoft Excel (Office 365, Microsoft corp., Washington, USA) and Stata version 16.1.^30^ Socio-demographic and practice characteristics of respondents were summarized using frequencies and percentages. Logistic regression was performed to identify HCP-associated predictors of a negative impact from the COVID-19 pandemic on immunization services. A negative impact on immunization practices was reported as a binary variable (impact or no impact) and defined as a modification in immunization practices or complete closure of the practice of a respondent who usually provides immunization to children. Covariates for the multivariable model were selected *a priori* and independent of univariate analyses, based on expert opinion and previous research documenting an association with on-time immunization coverage.^26,27^ Adjusted odds ratios (aOR) and 95% confidence intervals (CI) were calculated, and p-values <0.05 were considered statistically significant. Violin plots with medians and interquartile ranges (IQR) were generated using Python’s seaborn package to visualize changes in the proportion of in-person visits before versus during the pandemic. In respect to respondents’ qualitative input from the open-ended questions, thematic content analysis was performed by the first author. Respondents’ answers were clustered under emerging themes, and the number of comments within each theme was reported using frequencies and percentages.

## RESULTS

### Participant characteristics

A total of 475 responses were received from throughout Ontario, including 286 pediatricians (21.8% response rate) and 189 family physicians (9.5% response rate) within a 5-week period, from May 27 to July 3, 2020 (14.4% total response rate), a time frame that corresponded with the first wave of COVID-19 cases in Ontario. In total, 392 respondents (83%) reported usually providing vaccines to children and teenagers. In the pediatrician group, 239 (84%) were general pediatricians and 47 (16%) were pediatric subspecialists. From the subspecialist group, 21 respondents (44%) reported usually providing routine childhood immunization. In the family physician group, the vast majority reported usually seeing children as part of their practice (n=183, 97%) and providing vaccines to children (n=181, 96%), with children accounting for 26.2% of their patients on average (standard deviation of 16.8%). With respect to PPE, a majority of respondents had access to gloves (n=457, 96%), surgical masks (n=452, 95%), eye protection (n=399, 84%) and gowns (n=350, 74%), and about half of respondents reported having access to N95 respirators (n=220, 46%). Most respondents reported that their clinic was responsible for buying PPE (n=361, 76%), with fewer respondents receiving PPE from donations (n=78, 16%) or the government (n=65, 14%). All respondent characteristics are detailed in Table 1. A map of the province with an overview of the number of respondents by forward sortation area was also created (Figure 1). Of note, the initial 102 respondents from the GTA pediatrician listserv were not asked their practices’ forward sortation area and are therefore not represented on this map.

**Table 1:**
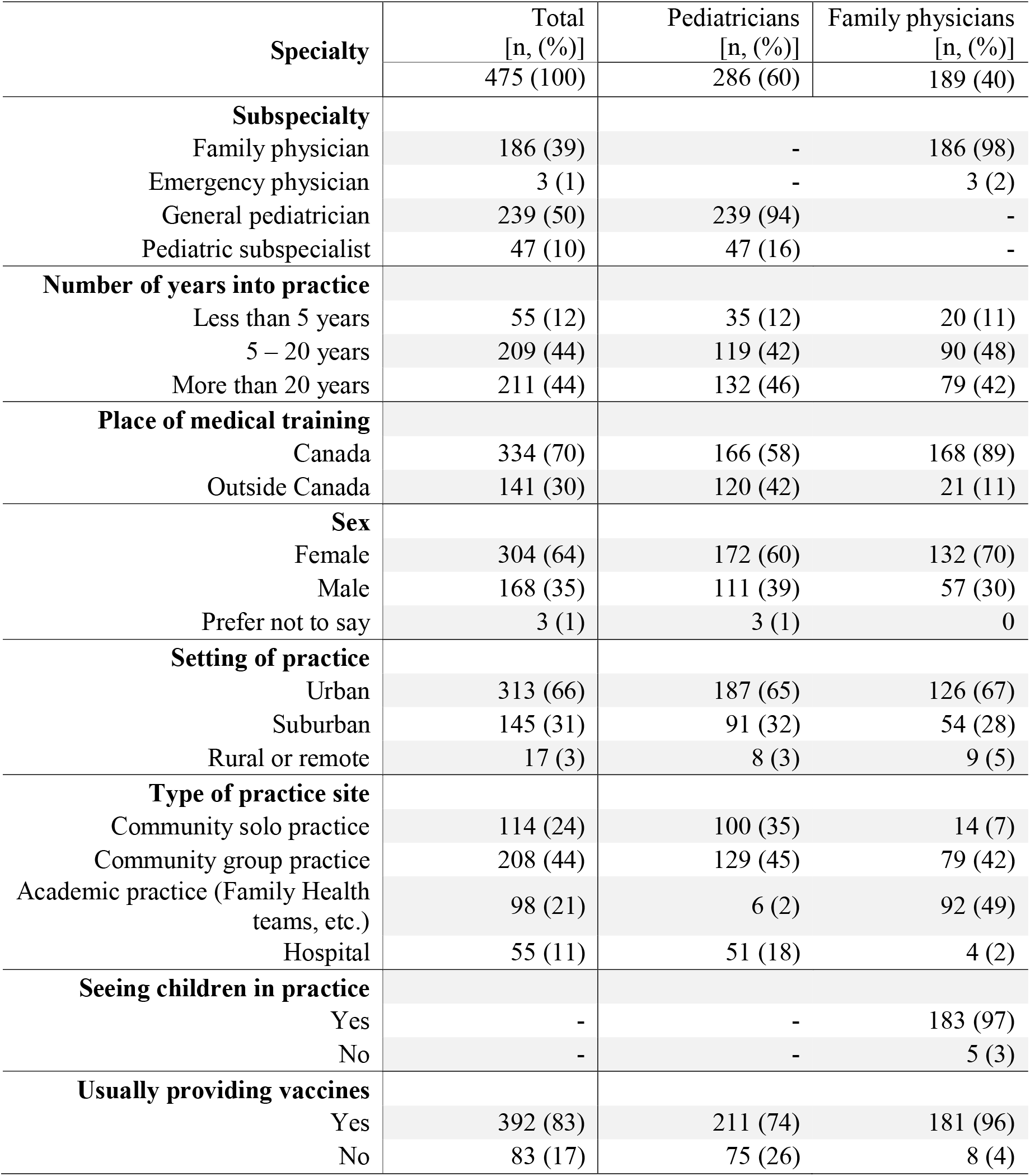
Characteristics of respondents.

| **Specialty** | **Total**<br>**[n, (%)]** | **Pediatricians**<br>**[n, (%)]** | **Family physicians**<br>**[n, (%)]** |
| --- | --- | --- | --- |
|  | 475 (100) | 286 (60) | 189 (40) |
| **Subspecialty** |  |  |  |
| Family physician | 186 (39) | - | 186 (98) |
| Emergency physician | 3 (1) | - | 3 (2) |
| General pediatrician | 239 (50) | 239 (94) | - |
| Pediatric subspecialist | 47 (10) | 47 (16) | - |
| **Number of years into practice** |  |  |  |
| Less than 5 years | 55 (12) | 35 (12) | 20 (11) |
| 5 – 20 years | 209 (44) | 119 (42) | 90 (48) |
| More than 20 years | 211 (44) | 132 (46) | 79 (42) |
| **Place of medical training** |  |  |  |
| Canada | 334 (70) | 166 (58) | 168 (89) |
| Outside Canada | 141 (30) | 120 (42) | 21 (11) |
| **Sex** |  |  |  |
| Female | 304 (64) | 172 (60) | 132 (70) |
| Male | 168 (35) | 111 (39) | 57 (30) |
| Prefer not to say | 3 (1) | 3 (1) | 0 |
| **Setting of practice** |  |  |  |
| Urban | 313 (66) | 187 (65) | 126 (67) |
| Suburban | 145 (31) | 91 (32) | 54 (28) |
| Rural or remote | 17 (3) | 8 (3) | 9 (5) |
| **Type of practice site** |  |  |  |
| Community solo practice | 114 (24) | 100 (35) | 14 (7) |
| Community group practice | 208 (44) | 129 (45) | 79 (42) |
| Academic practice (Family Health teams, etc.) | 98 (21) | 6 (2) | 92 (49) |
| Hospital | 55 (11) | 51 (18) | 4 (2) |
| **Seeing children in practice** |  |  |  |
| Yes | - | - | 183 (97) |
| No | - | - | 5 (3) |
| **Usually providing vaccines** |  |  |  |
| Yes | 392 (83) | 211 (74) | 181 (96) |
| No | 83 (17) | 75 (26) | 8 (4) |

**Figure 1:**
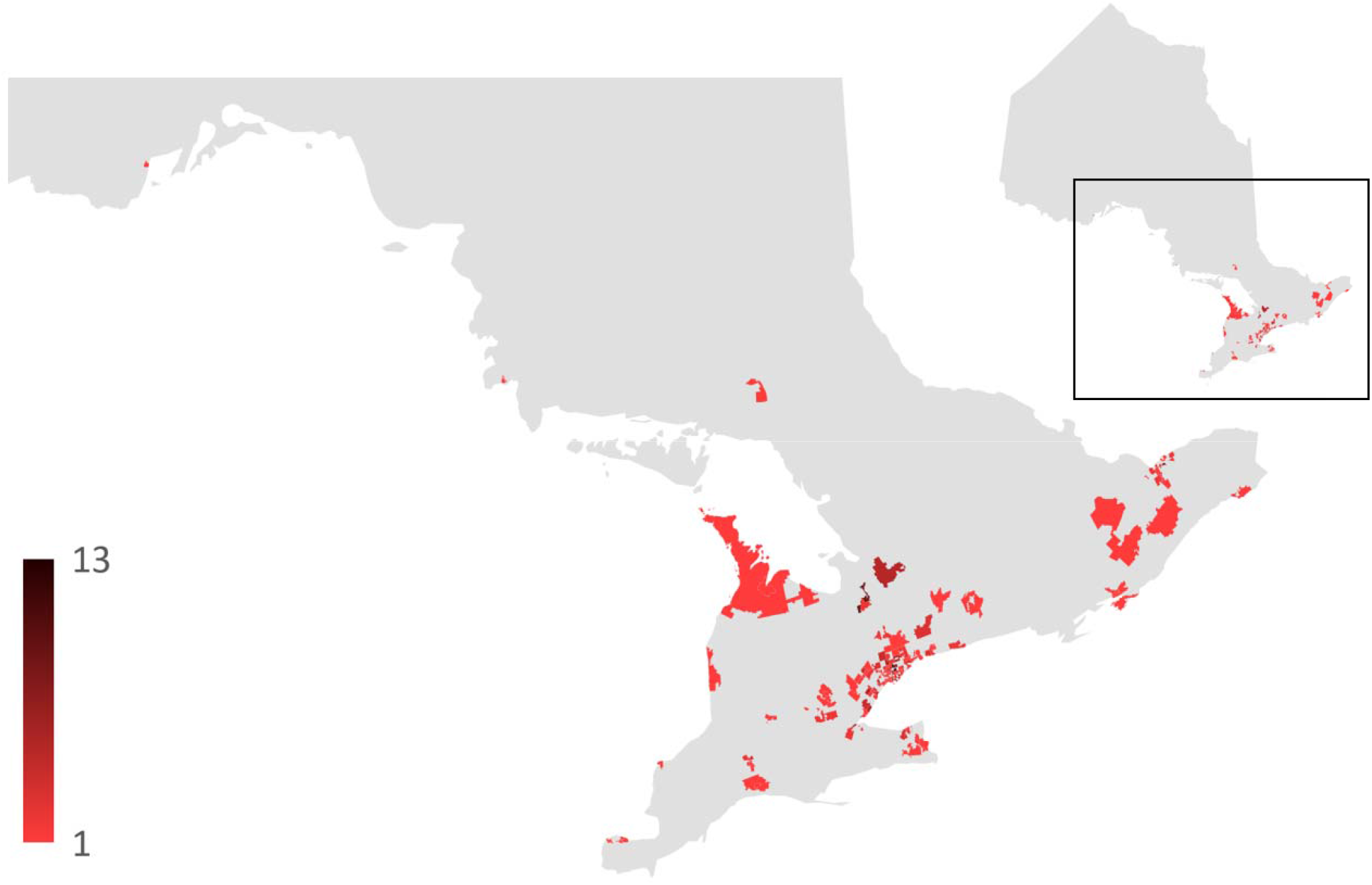
Number of survey respondents by forward sortation area.

### Impact on physicians’ practices

In total, 18 respondents (4%) reported that the COVID-19 pandemic caused a temporary closure of their practice, with the lack of PPE being the main reason for the closure (n=11, 61%). The median proportion of in-person visits reported by family physicians was 95.5% (IQR 90.0%-100.0%) before the pandemic and dropped to 12.0% (IQR 9.3%-20.0%) during the pandemic. Similarly, for pediatricians, the median proportion of in-person visits was 100.0% (IQR 95.0%-100.0%) before the pandemic and dropped to 25.0% (IQR 10.0%-50.0%) during the pandemic. The distribution of in-person visits before and during the pandemic is displayed in Figure 2.

**Figure 2:**
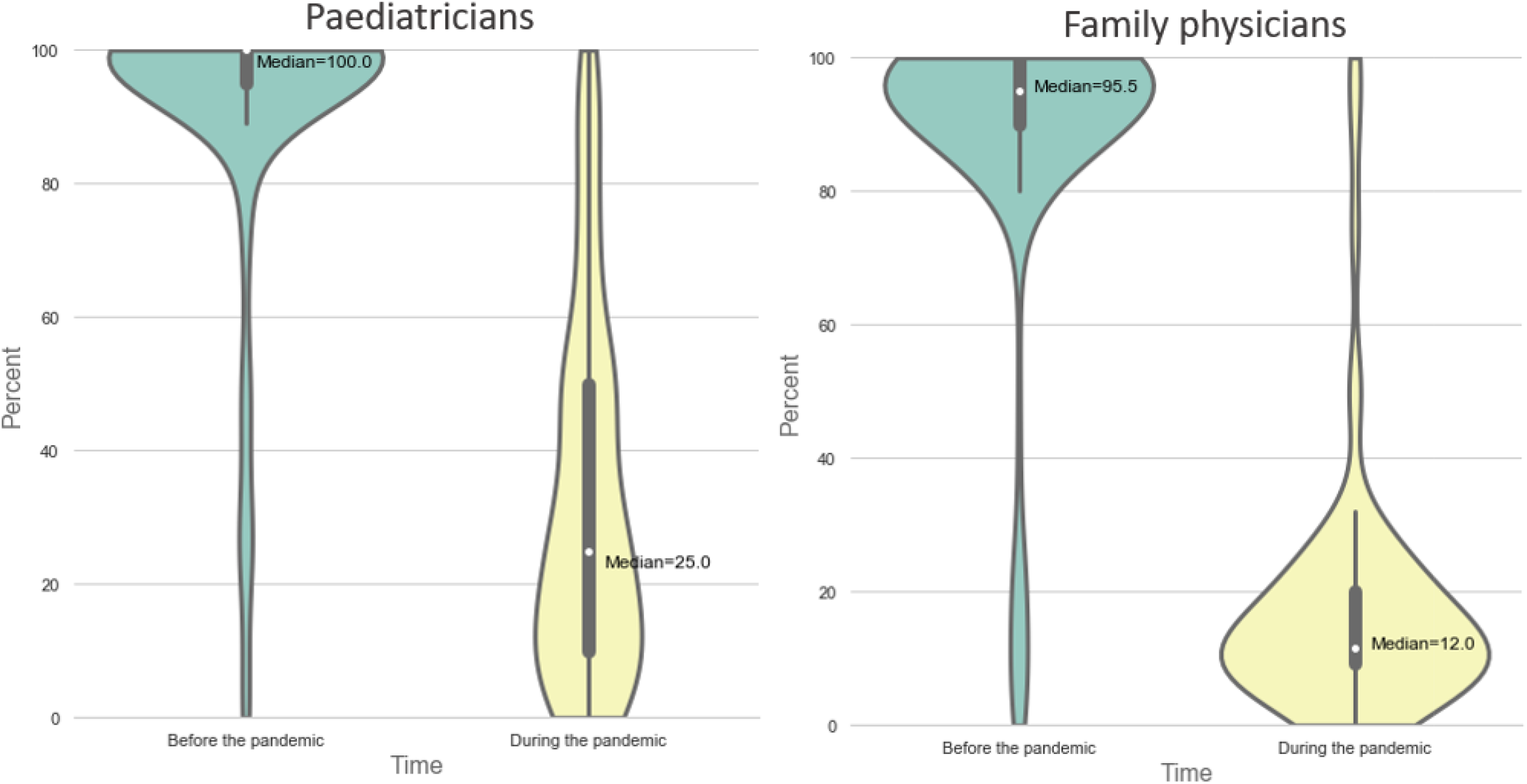
Violin plots of the proportion of in-person visits (%) befor e and during the pandemic for pediatricians and family physicians.

### Impact on physicians’ immunization services

In total, 175 (45%) of the 392 respondents who usually provide vaccination to children acknowledged a negative impact caused by the pandemic on their immunization services, including 18 respondents (5%) having closed their practice, 4 (1%) postponing all vaccines and 4 (1%) telling their patients to get vaccinated in another clinic. Most respondents reported only providing immunizations to children of a specific age (n=103; 26%), with the majority of this group continuing the vaccination of children aged 18 months and less (n=97, 94%), and postponing vaccination for children aged 4-6 years old (n=79, 77%) and teenagers (n=100, 97%). Fewer respondents reported only postponing specific vaccines (n=52, 13%), especially the rotavirus vaccine (n=33, 63%), the influenza vaccine (n=41, 79%), and the HPV vaccine (n=44, 85%). Figure 3 represents the reported impact of COVID-19 on immunization practices. In total, 118 respondents (30%) reported providing immunizations to patients from other clinics that were not offering this service anymore because of the pandemic, either frequently (n=19, 5%), occasionally (n=69, 18%) or rarely (n=30, 7%). A majority of respondents (n=271, 57%) reported that they did not have a system in place to keep track of their patients who may have missed vaccine doses. In respect to vaccines usually given in school settings (HBV, HPV and meningococcal quadrivalent vaccine), many of the survey participants reported not to provide these vaccines and leaving the missed doses to the schools when they would re-open (n=111, 28%) or to public health clinics (n=74, 19%).

**Figure 3:**
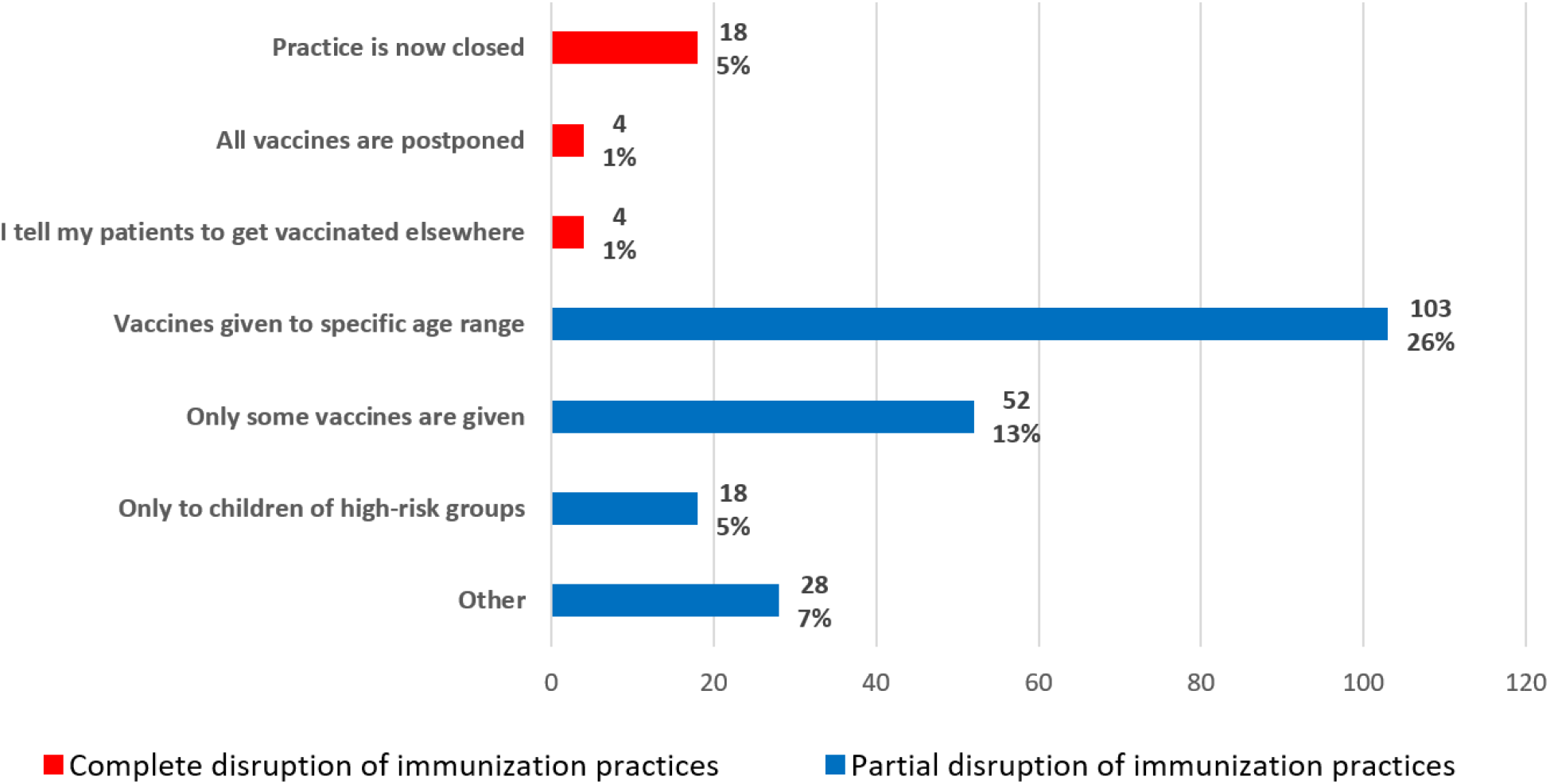
Bar chart representing the impact of COVID-19 on physicians’ immunization services for children. NB percentages are based on physicians who usually provide vaccines to children (n=392)

Most respondents were in disagreement or strong disagreement with the following statements: “I feel it is safe to postpone routine childhood immunizations because current physical distancing measures decrease the risk of vaccine preventable diseases” (n=430, 91%); “I feel it is preferable to postpone routine childhood immunizations to limit the spread of COVID-19” (n=434, 91%); and “I feel it is preferable to postpone routine childhood immunizations to limit the risk of COVID-19 exposure for healthcare workers” (n=419, 88%). A majority of respondents acknowledged that there were significant barriers to the delivery of immunization services pertaining to the COVID-19 pandemic (n=328, 84%). The most frequently identified barriers are shown in Figure 4. Figure 5 shows the PPE elements used by physicians to vaccinate asymptomatic children, as well as the elements that they think would be truly necessary.

**Figure 4:**
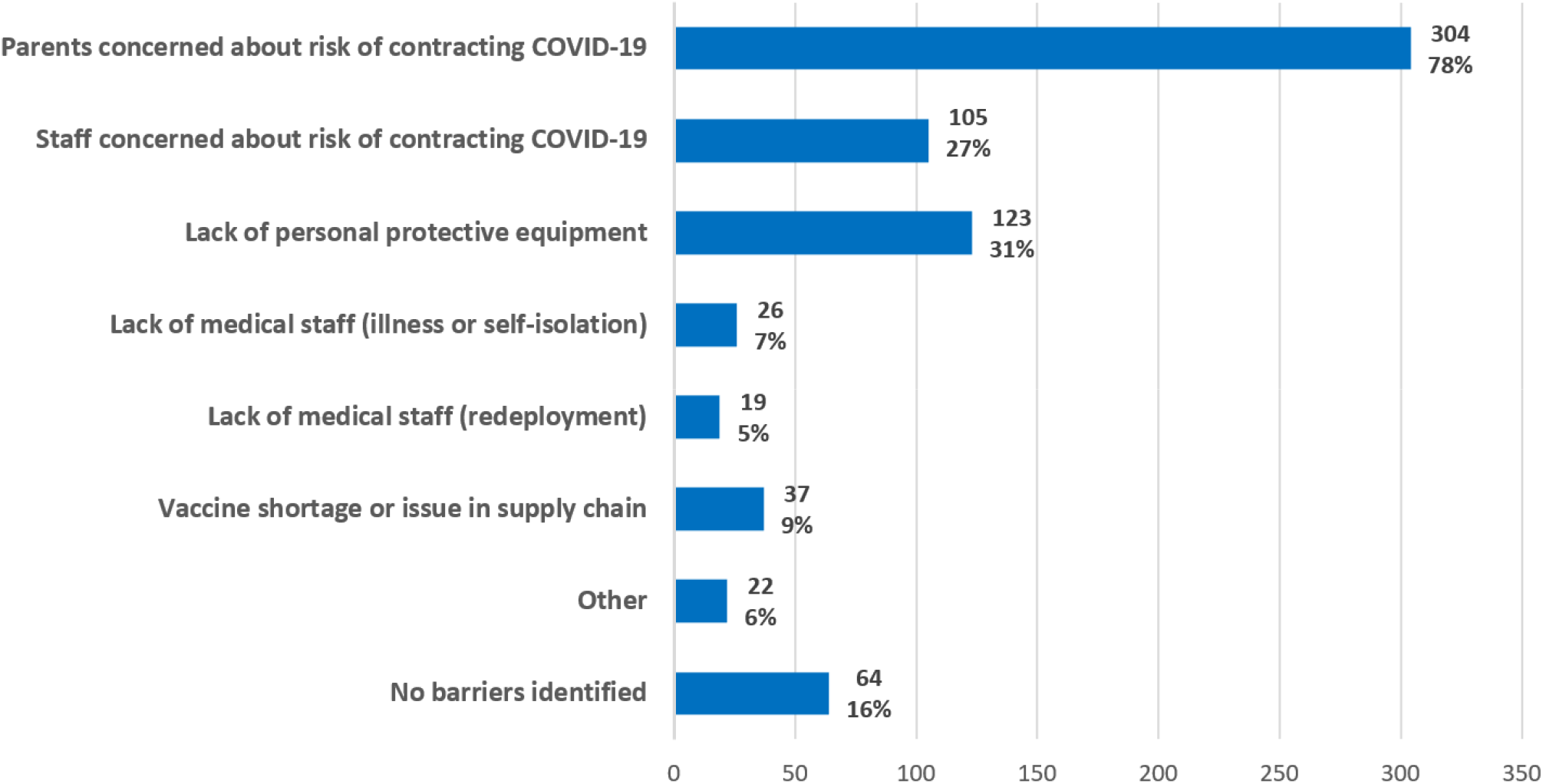
Bar chart representing the identified barriers to childhood routine immunization during the COVID-19 pandemic. NB percentages are based on physicians who usually provide vaccines to children (n=392)

**Figure 5:**
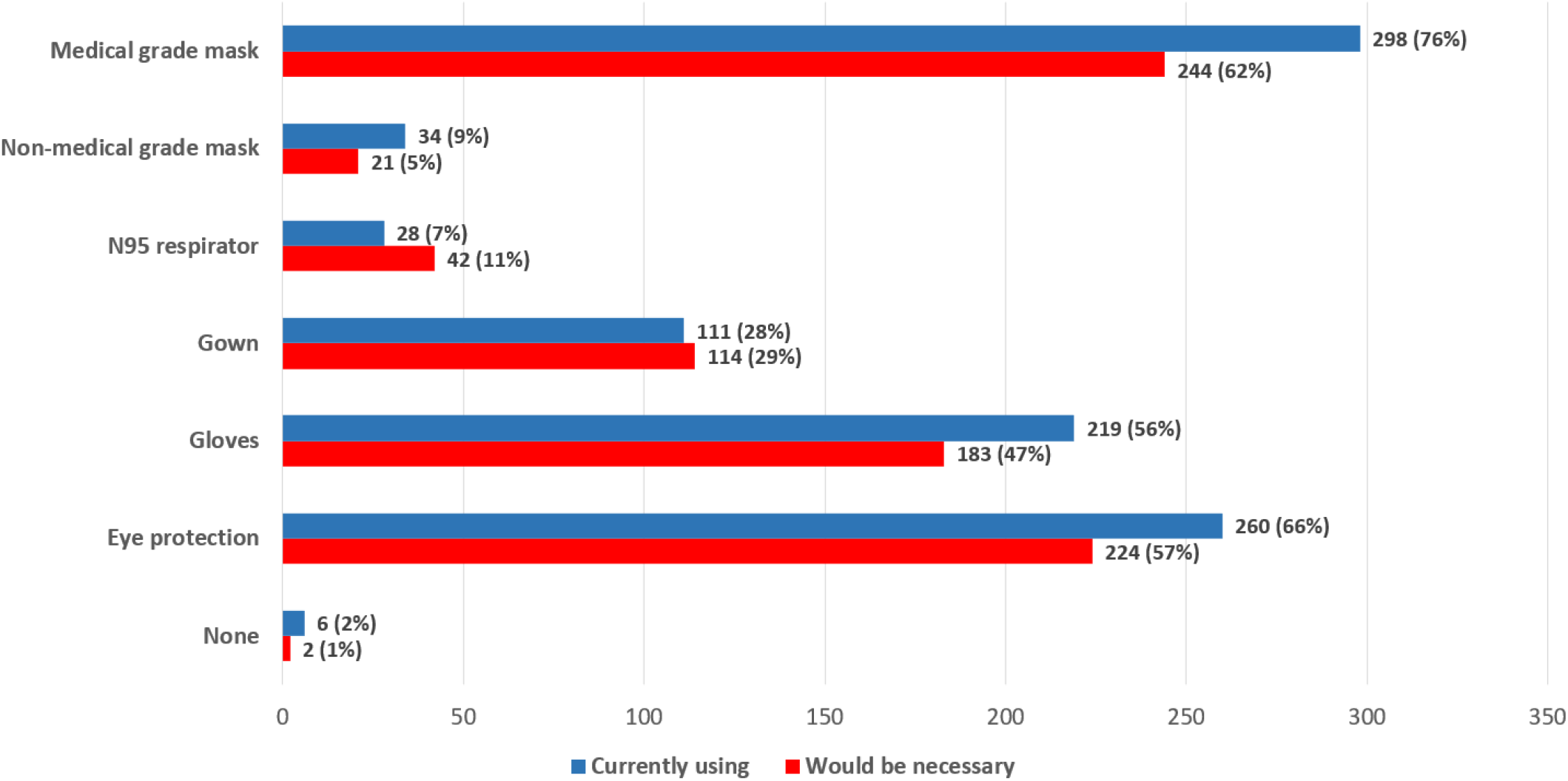
Bar chart representing the PPE elements currently used by HCP for vaccination of asymptomatic children, and which elements they think would be necessary. NB percentages are based on physicians who usually provide vaccines to children (n=392)

### Factors associated with decreased immunization services

In the multivariate logistic regression analysis, pediatricians were more likely to report that COVID-19 negatively impacted their delivery of immunizations than family physicians (aOR=2.08, 95% CI 1.31-3.30), while physicians based in suburban practices were less likely to report a negative impact than physicians in urban practices (aOR=0.61, 95% CI 0.39-0.97). Physicians in practice for less than five years were also more likely to report impacted immunization services compared to physicians in practice for 5-20 years (aOR=2.69, 95% CI 1.30-5.56). There was no statistically significant association between impacted immunization services and either physician’s sex or country of medical training. All associations, with odds ratio from the univariate analysis and adjusted odds ratio from the multivariate analysis are displayed in Table 2.

**Table 2:** Multivariable logistic regression of practitioner-level predictors of overall COVID-19 negative impact* on routine childhood immunization services.

| Characteristics | Univariate analysis |  | Multivariate analysis |  |
| --- | --- | --- | --- | --- |
|  | OR (95% CI) | p-value | aOR (95% CI) | p-value |
| **Specialty** |  |  |  |  |
| Family physician | ref | --- | ref | --- |
| Pediatrician | 1.73 (1.16-2.60) | **0.008** | 2.08 (1.31-3.30) | **0.002** |
| **Site of primary practice** |  |  |  |  |
| Community solo practice | ref | --- | --- | --- |
| Community group practice | 0.91 (0.56-1.48) | 0.714 | --- | --- |
| Academic practice<br>(Family health team) | 0.90 (0.51-1.60) | 0.726 | --- | --- |
| Hospital | 0.98 (0.28-3.42) | 0.973 | --- | --- |
| **Years of practice** |  |  |  |  |
| <5 years | 2.53 (1.24-5.16) | **0.011** | 2.69 (1.30-5.56) | **0.008** |
| 5-20 years | ref | --- | ref | --- |
| >20 years | 0.89 (0.58-1.36) | 0.589 | 0.75 (0.47-1.18) | 0.208 |
| **Physician's sex** |  |  |  |  |
| Female | ref | --- | ref | --- |
| Male | 1.06 (0.69-1.63) | 0.796 | 1.25 (0.80-1.97) | 0.327 |
| **Country of medical training** |  |  |  |  |
| Canada | ref | --- | ref | --- |
| Outside Canada | 1.06 (0.69-1.63) | 0.796 | 0.72 (0.44-1.18) | 0.192 |
| **Setting of primary practice** |  |  |  |  |
| Urban | ref | --- | ref | --- |
| Suburban | 0.65 (0.42-1.01) | 0.055 | 0.61 (0.39-0.97) | **0.037** |
| Rural or remote | 0.78 (0.24-2.53) | 0.682 | 1.10 (0.33-3.68) | 0.873 |
| **Practice in GTA** |  |  |  |  |
| No | ref | --- | --- | --- |
| Yes | 1.25 (0.83-1.89) | 0.283 | --- | --- |

\*Impact defined as a modification in physicians' immunization practices or complete closure of the practice. Responses of two physicians (0.5%) were excluded from the regression analyses due to missing data points.

### Thematic content analysis of open-ended questions

The first open-ended question from the survey pertained to further barriers identified by respondents regarding provision of childhood immunization services during the COVID-19 pandemic. In total, 137 (35%) of the 392 respondents usually administering vaccines provided answers to this question. The most frequently identified themes and barriers to immunizations was “school closures” (n=28, 20%), followed by “vaccine supply issues” (n=21, 15%) and “parental concerns” (n=20, 15%). Identified barriers from the respondents’ answers are listed in Table 3, with an associated representative quote. All survey respondents were also asked to provide suggestions to safely maintain childhood immunizations throughout the COVID-19 pandemic in a second open-ended question. In total, 189 (40%) of the total 475 respondents provided answers. The most frequent suggestions were: “assistance in providing personal protective equipment to practices” (n=65, 34%), “reorganization of patient flow” (n=48, 25%), “dedicated centers or practices for vaccination” (n=41, 22%), as well as “parental education and campaigning” (n=31, 16%). %). Identified suggestions from the respondents’ answers are listed in Table 4, with an associated representative quote.

**Table 3:** Barriers and impact to routine childhood immunization from respondents’ qualitative input.

| Barriers to immunizations | Number of related comments [n, (%)] | Illustrative comments |
| --- | --- | --- |
| School closures | 28 (20)* | School immunizations are impossible to keep up as no contact with public health about what is missed or started. Plus we do not have those immunizations in the office. |
| Vaccine supply issue | 21 (15) | It has brought the fragmentation of our system to the forefront- e.g. we have continued, schools haven't, families are confused. Public health reduced our vaccine quantities even though we maintained full service. This was an unnecessary barrier we had to expend time and energy to overcome. |
| Parental concerns | 20 (15) | Government puts such a fear into people of COVID-19 that parents can't see beyond one illness at present. |
| Lack of PPE | 17 (13) | Initially had minimal to no PPE with closure of clinic to virtual visits only, eventually bought PPE to offer immunizations prioritizing 18 months under. |
| Significant delays for older children | 16 (12) | We aren't able to bring in the older kids because we want to keep the office clean and safe for the newborns and infants. We have made this a priority because our group recognizes how important it is to vaccinate these children. |
| Reduced office hours | 14 (10) | Immunization may be delayed because parents do not want to come to the clinic and because I have limited the hours in my clinic |
| Unpreparedness | 14 (10) | At the start of the pandemic we didn't have a set way to deal with patients but as things evolved, we now have a smoother system. |
| Lack of support (Government, public health, etc.) | 11 (8) | I do not feel the government has supported physicians at all. As well public health continues to send the important message to families about vaccinating children yet they are not supporting offices either with respect to PPE or even stepping up to provide vaccines to older children (ages 4-6) or the grade 7 cohort. |
| Visits taking longer | 10 (7) | It takes me about 30-45 min per visit, because I disinfect the room between patients and have parents wait in the car. |
| Lack of public awareness / knowledge | 7 (5) | PPE and staff safety are the major concerns, as well as lack of education for parents regarding immunization and COVID 19 risks. |
| Concerns for staff safety | 7 (5) | Postponement may occasionally be necessary to protect me and staff if enquiry indicates that the parent(s), the patient, or house-hold members carry a risk of having Covid19 infection. |
| Practices' closure | 6 (4) | Our government agencies and medical associations were very slow from the start making it clear that many pediatric offices remained open and that immunizations were still important. Many GP offices in our community were shut down and remain so. There was a local assumption by many that we were as well. |
| **Total number of comments** | **137 (35)\*\*** |  |

\*Percentages were calculated based on number of free text comments received for this question.
\*\*Percentage was calculated based on the number of respondents who usually provide vaccines for children and were presented with this question

**Table 4:** Suggestions from respondents to safely maintain immunization coverage for children throughout the COVID-19 pandemic.

| Possible solutions | Number of related comments [n, (%)] | Illustrative comments |
| --- | --- | --- |
| Assistance in providing PPE to practices | 65 (34)* | I have written letters to local businesses, the government and the OMA requesting assistance with obtaining PPE and have received no assistance or replies. Our clinic is supporting several other clinics in the area by providing pediatric care for their patients when other offices close. Without PPE, our office will also have to close. |
| Reorganising patient flow | 48 (25) | One parent per child. Parents are called the previous day by me and I go through the child's development and answer all their questions. The following day is then only for the actual vaccination. It works about 85% of the time. |
| Dedicated centers or practices for vaccination | 41 (22) | Trying to establish a community hospital site for routine vaccinations to make sure that primary care providers with closed offices have a simple alternative. I would love to see all childhood immunizations given at a centralized location and NOT in pediatrician's offices. |
| Education campaigns targeting parents | 31 (16) | The phone calls really help. We actively call parents and explain our precautions, and that we feel strongly that immunization should not be delayed. |
| Patient screenings | 28 (15) | The patients appreciate when we screen them over the phone and go through the protocol used, bring reassurance that we are taking the appropriate measures. |
| Centralized guidance | 14 (7) | Better guidance from groups like the CPS which has been virtually completely lacking during this pandemic. In addition, guidance for the next wave of COVID-19, and for the next pandemic, both of which will come. |
| Masking of patients and parents | 12 (6) | Mom and dads must be masked when they come in, distancing, plexiglass at our reception. |
| Vaccines given outside of practices (e.g. drive-thru) | 9 (5) | We offer vaccine administration in the parking lot where they can wait in their car after the shot to ensure no reaction. This option is appealing to most parents. |
| Subsidies for practices | 7 (4) | We should reimburse pediatricians offices for the extra time and cost encountered by them to provide such service. I suggest adding Covid surcharge in amount of 30% to all immunization related visits |
| Ensure adequate vaccine supply | 7 (4) | A centralized provincial clearinghouse for ordering vaccines and mailing of vaccines directly to our offices would decrease likelihood of shortages and delays in vaccine administration. |
| Simplify vaccine schedule | 6 (3) | Give varicella and Pediacel during the same visit for 15-18 months old instead of 2 separate office visits. Give the 4-6 year-old vaccines at the same time as flu vaccine. |
| Centralized immunization record | 4 (2) | Centralize the immunization data (should have already been done years ago, would be really helpful about now), so everybody knows accurately where they stand re: vaccines. Nobody should have to call a doctor's office to "find out" if their vaccines are up to date. |
| **Total number of comments** | **189 (40)\*\*** |  |

\*Percentages were calculated based on number of free text comments received for this question.
\*\*Percentage was calculated based on the total of survey respondents.

## DISCUSSION

To our knowledge, this is the first study to evaluate and quantify the impact of the COVID-19 pandemic on childhood immunization practices in Canada and to identify the possible practitioner-level predictors of this impact. Our findings can inform the implementation of policies to mitigate a decrease in immunization coverage throughout future waves of the COVID-19 pandemic and avoid a potential surge of VPDs later on.^31,32^

The indirect impacts of the COVID-19 pandemic on access to medical services, including routine immunizations, represent an important burden on children’s health that may outweigh the burden of the infection itself.^15,17,18^ The findings from our study highlight this reality: close to half of the participating physicians reported a negative impact of the COVID-19 pandemic on their immunization services, with a few HCP even postponing all of their patients’ vaccines. Most respondents reported that parental concerns around COVID-19 were the most significant impediment to appropriate immunization coverage of children; our qualitative analysis also supported this finding. This barrier should be urgently addressed, and public health units and governmental bodies should invest in education campaigns for parents with a focus on vaccine safety, effectiveness, and healthcare-seeking behaviors. Parents and patients should be informed that measures aiming to mitigate the transmission of COVID-19 in healthcare settings have been implemented. Further studies exploring how COVID-19 has influenced parents’ perception of vaccines are needed in order to appropriately target their concerns and limit a decrease in vaccine coverage. These studies should also explore parental perspectives of COVID-19 vaccines, and how this could influence their acceptance of other routine childhood immunizations.^33^

Over a quarter of the survey’s respondents mentioned providing vaccines to patients who are not usually rostered to their clinic, which may suggest that the number of closed practices are under-represented in our study. This finding underlines the challenges that some parents may face in accessing vaccines for their children during the COVID-19 pandemic, as they have to navigate the healthcare system to find clinics that remain open and would be willing to provide vaccines to their children. Given that there is no central electronic repository of immunization records in most Canadian provinces, having timely and precise information on patients’ vaccine uptake is challenging.^39^ Accordingly, a majority of our survey’s respondents acknowledged that they do not have a system in place allowing them to keep track of patients that may have missed immunization doses. This may introduce significant challenges in planning catch-up immunization schedules in a systematic way including prioritization of specific vaccines or higher risk patients. Moreover, patients seeking catch-up vaccines will add to increased volumes on the healthcare system during pandemic recovery phases, which further highlights the importance of maintaining up-to-date uptake of routine immunizations throughout the pandemic. Practices that are closed or do not have the capacity to provide immunization services for all children should therefore make sure that their patients have been referred to other practices to receive their vaccines. In our study, many respondents also suggested the creation of clinics specifically designed to provide vaccines to children to facilitate this process. Some public health units in Ontario have successfully implemented a similar system, which may be scaled up in other settings.^34^ Most respondents also highlighted the importance of PPE to continue providing vaccines safely and expressed concern about access. In the interim guidance document from the Public Health Agency of Canada (PHAC) for outpatient and ambulatory care during COVID-19, it is recommended that staff should wear a medical mask at all times and should strongly consider the use of eye protection (e.g. face shields) when providing direct patient care.^12^ This recommendation is aligned with the answers that we received in our survey, in which a majority of respondents reported using masks with or without eye protection to administer vaccines to asymptomatic children.

Our study also found that the immunization services provided by pediatricians, as well as HCP from urban settings and the ones who have been in practice for a shorter period of time, were significantly more impacted by the pandemic. While this may inform which HCP should be preferentially supported in their efforts to continue providing immunizations to children, our study did not explore the possible reasons behind these findings. In Ontario, given that a substantial proportion of COVID-19 cases have been diagnosed in populous cities, it is not surprising that urban settings would be more impacted than suburban or rural settings.^35^ On the other hand, the reasons for preferential negative impact on pediatricians and early-career physicians immunisation practice are unknown. Previous studies have found that more recently graduated physicians had decreased odds of believing vaccines are efficacious and safe compared to graduates from a previous 5-year period, and it is possible that some recent graduates did not prioritise immunisation visits as much as their more experienced colleagues did. It is also possible that early-career physicians had been redeployed to other clinical tasks related to the COVID-19 pandemic more frequently than their more senior colleagues, which could have impacted their immunization practices.^36^

Many of our study’s respondents have reported maintaining vaccines in patients aged ≤ 18 months, which is in keeping with PHAC recommendations to prioritize primary immunization series during COVID-19.^12^ Most of our study’s participants have been postponing vaccines for older children, including those usually given in school settings, which they would leave to the schools when they would re-open. Given the occurrence of a second and third waves of the pandemic in Ontario, most public health units have cancelled their school-based immunization program for the 2020-2021 school year.^37^ Public health units have therefore recommended that students should instead receive these vaccines through their primary care provider. The fact that healthcare providers’ have reported decreased immunization practices and clinical activities during the first wave of the pandemic suggests that they may not have the capacity to catch-up on immunizations usually given in school settings for all of their patients. Repeated surveys of healthcare providers during subsequent waves of the pandemic should be conducted, as recommendations on vaccination programs from public health units and government bodies are changing based on the COVID-19 epidemiological situation.

Our study has several limitations. First, the low response rate and the potential introduction of selection bias due to the convenience sampling of respondents can limit the external validity and generalizability of our study results. Additionally, practice size and the number of children seen in each practice was not collected from the survey respondents, which limits the quantification of the impact on vaccine coverage at a population-level. Also, while physicians represent a substantial proportion of the HCP providing vaccines to children in Ontario, we have not evaluated the impact on the immunization services provided by nurse practitioners, nurses, and pharmacists. Lastly, in view of the cross-sectional nature of the study, we were only able to capture the immunization practices of physicians within a limited timeframe and during the first wave of the COVID-19 pandemic, well before vaccines against COVID-19 were available.

## CONCLUSION

Our survey study reports that COVID-19 has caused substantial modifications to physicians’ pediatric immunization services across Ontario in the initial wave of the pandemic, which suggests that there could be a cohort of under-vaccinated children at increased risk of VPDs when public health measures aiming to mitigate the transmission of COVID-19 are relaxed. The survey respondents have identified multiple barriers to routine childhood immunizations, with parents’ concerns about their children contracting COVID-19 in healthcare settings being the most significant. Concerted efforts by HCP, public health units and government bodies are required to ensure appropriate provision of routine childhood immunizations throughout the COVID-19 pandemic, catch-up on any missed vaccines, and educate parents and caregivers about the importance of maintaining vaccine coverage for their children.

## Data Availability

The survey answers were collected using Research Electronic Data Capture (REDCap) software. Survey participantscould decide not to opt in for the incentive draw if they wished to keep their responses completely anonymous. The collected emails and names were not linked with the participants' responses to maintain confidentiality.

## ABBREVIATIONS

aOR: Adjusted Odds Ratio
CDC: Centers for Disease Control
CI: Confidence Interval
COVID-19: Coronavirus Disease 2019
GTA: Greater Toronto Area
HPV: Human Papillomavirus
IQR: Interquartile range
PPE: Personal Protective Equipment
REDCap: Research Electronic Data Capture
PHAC: Public Health Agency of Canada
US: United States
VPDs: Vaccine Preventable Diseases

## ACKNOWLEDGEMENT

We would like to acknowledge all pediatricians and family physicians who participated in this survey study. We would also like to thank Robyn Neville-Kett from the Pediatricians’ Alliance of Ontario and the Department of Family and Community Medicine at the University of Toronto for their help with the distribution of the survey.

